# ALARM-Net: Knowledge Graph-Informed, Onset-Aware Multichannel EEG Seizure Detection with Event-Level False-Alarm Suppression

**DOI:** 10.64898/2026.05.17.26353436

**Authors:** Osman Yildiz, Abdulhamit Subasi

## Abstract

Automated EEG seizure detection is clinically constrained by the event-level false-alarm burden of long-term recordings and by detectors that act as opaque window classifiers without clinically interpretable spatial reasoning. We propose ALARM-Net (Alarm-Level Adaptive Rejection Module), a knowledge-graph-informed, onset-aware seizure detection framework with event-level false-alarm suppression, evaluated on the Temple University Hospital Seizure Corpus (TUSZ) v2.0.6. We first construct a clinically grounded EEG knowledge graph over bipolar-montage channels, cortical regions, hemispheres, anatomical adjacency, contralateral homology, and focal-onset priors. The knowledge graph is used not as a post-hoc label map but as a computational substrate for the detector: it defines the graph-attention topology, supplies per-channel clinical priors, and supports an auxiliary onset-channel head trained to recover expert-annotated seizure-onset channels. The resulting detector preserves window-level detection (AUROC 0.94) while localizing seizure onset on 19 held-out patients (subject-level AUC 0.72–0.77; 18–19 of 19 patients above chance; Wilcoxon p < 10−5); independently, its emergent channel attention — obtained without onset supervision — aligns with expert-annotated onset channels (17 of 19 subjects; p < 0.001). For the event-level evaluation, a knowledge-graph-informed heterogeneous GNN suppressor (KGSuppressor) scores alarm proposals using per-channel detector embeddings organized by the EEG knowledge graph, reducing false alarms per 24 h by 47.3% on the held-out evaluation split (19.55 → 10.30) at a dev-selected operating point while preserving nearly all seizure sensitivity (strict sensitivity cost of 0.8 percentage points), outperforming a gradient-boosting baseline that requires 7.8 percentage points of sensitivity loss for comparable FA reduction. By grounding multichannel EEG evidence in clinical anatomy and converting it into clinically usable alarms, ALARM-Net supports the feasibility of knowledge-graph-grounded, onset-aware analysis as an interpretable layer for EEG seizure detection.

## 1. Introduction

Continuous electroencephalography (EEG) monitoring is the standard approach for detecting seizures in patients with epilepsy and those at high seizure risk in critical-care settings. Automated seizure detection systems assist clinicians by identifying potential seizure events for review, operating within an alarm-driven workflow in which clinical decisions are based on generated alarms rather than window-level probability outputs [3,5]. Consequently, the practical value of these systems depends not only on seizure detection accuracy but also on their ability to minimize false alarms while maintaining high sensitivity. Although advances in deep learning have significantly improved EEG seizure detection, excessive false alarms remain a major barrier to clinical adoption, contributing to alarm fatigue, reduced clinician trust, delayed responses, and workflow disruptions. This challenge is particularly pronounced in multichannel EEG recordings, where artifacts, physiological noise, electrode disturbances, and non-epileptic rhythmic activity frequently mimic seizure patterns.

Most existing seizure detection methods focus on window-level classification using architectures such as CNNs, RNNs, transformers, and hybrid deep learning models. While effective for segment-level discrimination, these approaches often fail to fully capture the complex spatial-temporal dependencies and contextual relationships present across EEG channels and generally lack explicit incorporation of clinical knowledge regarding electrode connectivity, anatomical organization, seizure propagation, and alarm evolution. Recent advances in Knowledge Graphs (KGs) and Graph Neural Networks (GNNs) provide a promising alternative by enabling structured representation and reasoning over clinically meaningful relationships among EEG channels, cortical regions, seizure-onset characteristics, and temporal event dynamics. By integrating domain knowledge with graph-based learning, these approaches facilitate anatomically grounded spatial reasoning, improved modeling of seizure propagation and alarm behavior, and enhanced interpretability, thereby offering a more clinically meaningful framework for multichannel EEG seizure detection.

ALARM-Net (Alarm-Level Adaptive Rejection Module) is a knowledge-graph-guided, onset-aware framework for multichannel EEG seizure detection that integrates event-level false-alarm suppression on the TUSZ v2.0.6 dataset. Unlike conventional approaches that focus solely on optimizing seizure detectors, ALARM-Net adopts a modular architecture in which a fixed upstream detector generates seizure probability trajectories, while event-level alarm decisions are refined through KGSuppressor, a heterogeneous graph neural network operating on channel-level detector embeddings organized by a clinically grounded EEG knowledge graph. This graph encodes anatomical, hemispheric, and clinical relationships among EEG channels, cortical regions, and seizure-onset characteristics, enabling relational reasoning for distinguishing true seizure events from false alarms.

At the detection stage, ALARM-Net employs a knowledge-graph-informed graph attention network that incorporates clinically derived channel connectivity and seizure-onset priors. An auxiliary onset-localization branch further identifies seizure-onset channels, providing anatomically meaningful interpretation of seizure generation and propagation. By combining knowledge-driven spatial reasoning with event-level alarm modeling, ALARM-Net enhances interpretability through alignment with clinically annotated seizure-onset regions while substantially reducing false alarms and maintaining clinically acceptable seizure sensitivity. This integrated framework improves the robustness, trustworthiness, and practical clinical applicability of automated EEG seizure detection systems.

Although EEG seizure monitoring is fundamentally alarm-oriented, most existing approaches continue to emphasize window-level evaluation metrics such as AUROC, AUPRC, and F1 score [3,14,15]. While these measures assess classification performance, they do not necessarily reflect the clinical usefulness of a system when predictions are translated into seizure alarms. Recent studies have shown that models reporting low false-alarm rates on curated datasets may exhibit substantially higher false-alarm burdens when evaluated on larger, more heterogeneous datasets using standardized event-level evaluation protocols [3,4], highlighting the gap between statistical performance and real-world clinical utility. The Temple University Hospital Seizure Corpus (TUSZ) v2.0.6 [1,2], one of the largest and most clinically diverse EEG seizure datasets, has been widely used to evaluate graph-based, transformer-based, and foundation-model approaches [6,7,15]. However, differences in preprocessing, dataset versions, and evaluation methodologies continue to hinder direct comparison across studies.

Motivated by these challenges, this work investigates the event-level false-alarm behavior of a high-performing seizure detector on TUSZ v2.0.6 and reveals that false alarms are disproportionately concentrated among a small subset of patients. To mitigate this burden, the study introduces ALARM-Net, a knowledge-graph-informed framework that performs event-level false-alarm suppression through KGSuppressor, a heterogeneous graph neural network that exploits EEG knowledge graph relationships and channel-level detector embeddings. By incorporating clinically motivated alarm reasoning and suppression strategies, ALARM-Net enhances the reliability, interpretability, and practical clinical utility of automated EEG seizure detection systems while addressing one of the major barriers to real-world deployment.

The main contributions of the work are fivefold: (1) the construction of a clinically grounded EEG knowledge graph incorporating bipolar EEG channels, cortical regions, hemispheres, anatomical adjacency, contralateral homology, and seizure-onset priors; (2) the development of a knowledge-graph-informed, onset-aware graph attention detector with an auxiliary onset-localization branch; (3) validation of the detector’s clinical grounding through both supervised onset-channel prediction and post-hoc alignment of learned attention with expert-annotated seizure-onset regions; (4) the introduction of KGSuppressor, a knowledge-graph-informed event-level false-alarm suppression module; and (5) comprehensive event-level evaluation and ablation studies demonstrating that graph-based suppression improves alarm usability while preserving detection performance and enhancing interpretability. Together, these contributions establish a clinically interpretable, knowledge-graph-driven framework that bridges multichannel EEG analysis and clinically actionable seizure alarms.

The remainder of this paper is structured as follows. Section 2 provides an overview of related work in EEG seizure detection, false-alarm suppression, and graph-based learning approaches. Section 3 describes the dataset, preprocessing procedures, and the baseline seizure detection model. Section 4 presents the proposed ALARM-Net framework, including the knowledge-graph-informed detector and the KGSuppressor module. Section 5 reports the experimental results, comparative analyses, and ablation studies. Section 6 discusses the findings and their clinical implications. Section 7 outlines the study limitations and future research directions. Finally, Section 8 summarizes the main conclusions and contributions of the work.

## 2. Related Work

Research on window-level seizure detection has evolved through multiple generations of methodologies. Initial approaches relied on manually engineered spectral, temporal, and nonlinear EEG features, including band power, line length, and sample entropy, which were subsequently classified using conventional machine learning algorithms such as Support Vector Machines (SVM), Random Forests, and XGBoost [11,12]. The emergence of convolutional neural networks (CNNs) shifted the field toward end-to-end learning by reducing dependence on handcrafted feature extraction [14]. More recently, graph-based approaches have modeled EEG channels as interconnected nodes within a graph structure, allowing Graph Neural Networks (GNNs) to capture spatial relationships among channels and achieve strong window-level detection performance on TUSZ [8,13,15]. This progression has further expanded with the development of EEG foundation models [6,7], which leverage large-scale pretraining strategies. Nevertheless, meaningful comparison between foundation-model and graph-based approaches remains challenging because studies often differ in dataset versions, preprocessing procedures, experimental settings, and evaluation criteria.

While window-level metrics remain widely reported, recent efforts have highlighted the importance of evaluating seizure detection systems using clinically meaningful event-level measures. The SzCORE 2025 framework [3,4] formalized key event-level metrics, including false alarms per 24 hours (FA/24h), strict and reachable F1 scores, and seizure onset-detection latency, explicitly distinguishing between window-level metrics that characterize statistical discrimination and event-level metrics that better reflect clinical utility. This framework emphasizes that clinician-facing performance should be assessed primarily through event-level outcomes rather than segment-level predictions. For seizure-event matching on TUSZ, the Temple University Hospital NEDC scoring framework [5] serves as a widely adopted reference standard. Following common practice, we employ a one-to-one event matching strategy with a 30-second temporal collar to account for uncertainties in seizure onset annotations.

Several strategies have been proposed to mitigate false alarms in automated seizure detection systems. Existing approaches generally fall into two categories: post-processing techniques that refine detector outputs through probability smoothing, temporal filtering, or hysteresis-based thresholding [3,15], and ensemble-based methods that combine predictions from multiple detectors to improve robustness [4]. ALARM-Net complements these approaches by introducing a dedicated event-level suppression mechanism that operates independently of the underlying seizure detector. Rather than modifying detector predictions directly, the framework evaluates alarm proposals using graph-structured representations derived from per-channel detector embeddings and organized according to a clinically informed EEG knowledge graph. Conventional probability- and morphology-based alarm descriptors are retained as baseline features, while the proposed graph-based suppressor leverages relational information across EEG channels to improve discrimination between true and false alarms. Because the framework only requires access to channel-level detector embeddings, it can be integrated with a broad range of multichannel seizure detection architectures that provide channel-specific representations.

## 3. Data and Base Detector

### 3.1. Dataset

This study utilizes the Temple University Hospital Seizure Corpus (TUSZ) v2.0.6 [1,2], one of the largest publicly available clinical EEG datasets for seizure detection research. Following the official subject-disjoint train/dev/eval partition, recordings were filtered to include only those with the complete TCP-AR montage and a minimum duration of 60 seconds, resulting in 5,184 recordings: 3,199 for training, 1,194 for development, and 791 for evaluation. These recordings correspond to 296 training subjects, 41 development subjects, and 41 evaluation subjects, with no overlap between subject groups. Ground-truth seizure annotations extracted from the *csv_bi* files yielded 321 seizure events in the development set and 360 events in the evaluation set, with neighboring seizure segments separated by less than one second merged into single events to reduce annotation fragmentation. The evaluation partition was maintained as a strictly held-out test set and was not used for threshold selection, hyperparameter optimization, feature engineering, or model development. All operating points and model-selection decisions were determined exclusively on the development set and subsequently applied to the evaluation set without modification.

### 3.2. Preprocessing

The raw EEG recordings are first transformed into the standard 18-channel TUH TCP-AR bipolar montage and resampled to 200 Hz. The signals are then divided into overlapping 60-second windows with a 15-second stride, providing temporal continuity between adjacent segments. For model training and inference, consecutive windows are grouped into chunks of 20 windows, corresponding to approximately 5 minutes of EEG data per chunk. This chunk-based processing strategy enables the model to capture longer temporal dependencies but also introduces a limitation whereby recordings shorter than five minutes cannot be processed and are therefore excluded from model outputs.

### 3.3. Run A: Base Detector

The baseline seizure detector, denoted as Run A, is a window-level classification model comprising approximately 470K trainable parameters. The architecture integrates four principal modules: (1) a Temporal Convolutional Network (TCN) employing dilated one-dimensional convolutions to extract temporal features independently from each EEG channel [9]; (2) a Graph Attention Network (GAT) that models spatial interactions among the 18 EEG channels by representing them as nodes within a graph and learning attention-based inter-channel relationships [8]; (3) a quality-aware gating mechanism, which estimates signal quality using measures such as line-noise power, amplitude clipping, and the proportion of flat segments, thereby reducing the influence of poor-quality windows during training and aggregation; and (4) a graph-based regularization strategy that promotes temporal consistency between neighboring window predictions.

For each 60-second EEG segment, Run A generates a seizure probability score ranging from 0 to 1. On the development set, the model achieves an AUROC of 0.937, an AUPRC of 0.275, and a median subject-level AUROC of 0.956, indicating robust window-level classification performance. Nevertheless, direct comparisons with previously reported TUSZ results should be interpreted cautiously because of variations in dataset releases, preprocessing pipelines, and evaluation methodologies.

The model is trained for 30 epochs using the Adam optimizer with a learning rate of 3 × 10^−4^ and weight decay of 1 × 10^−4^. Optimization employs a cosine annealing learning-rate schedule (T_max = 30, minimum learning rate = 1 × 10^−5^), gradient clipping with a maximum norm of 1.0, and early stopping based on development-set AUROC with a patience of seven epochs. Each training step processes four EEG chunks with internal sub-batches of 80 windows and maintains a target positive-window ratio of 30%. The objective function consists of weighted binary cross-entropy (positive-class weight = 4.0) combined with three regularization components: a persistence penalty (λ = 2.0) that discourages unstable predictions, a spatial smoothness constraint over the channel graph (λ = 5.0), and a temporal smoothness constraint across adjacent windows (λ = 5.0). All experiments are conducted using a fixed random seed of 42, and no seed averaging is performed.

### 3.4. Window-Level vs Event-Level Evaluation

We differentiate between window-level and event-level evaluation paradigms. Window-level metrics, including AUROC, AUPRC, and segment-level F1 score, consider each 60-second EEG window as an independent classification instance. In contrast, event-level metrics assess the correspondence between temporally continuous predicted seizure events and annotated ground-truth seizure events through a one-to-one matching strategy. Since clinical decision-making is driven by alarms representing seizure events rather than individual window probabilities, event-level evaluation provides a more realistic measure of clinical utility. Consequently, following the recommendations of the SzCORE 2025 framework, we adopt strict F1 score, strict sensitivity, precision, and false alarms per 24 hours (FA/24h) as the primary performance metrics for assessing clinical applicability.

### 3.5. Strict Reporting Convention

Since Run A processes EEG data in fixed 5-minute chunks, recordings shorter than five minutes do not generate model predictions. As a result, certain annotated seizure events may occur within recordings that are excluded from processing and therefore remain undetectable by the model. To ensure a rigorous and unbiased assessment, all primary event-level performance metrics are calculated using strict evaluation denominators, whereby every annotated ground-truth seizure event is included in the analysis and any event occurring in an unprocessed recording is counted as a missed detection. This conservative evaluation strategy prevents artificially inflated performance estimates that could arise from excluding unreachable events through reachability-adjusted denominators. For completeness and additional interpretation, results based on reachability-adjusted evaluation are presented and discussed in Section 6.

### 3.6. EEG Knowledge Graph

The proposed detector is built upon a clinically informed EEG knowledge graph represented as a heterogeneous graph structure. The graph incorporates three categories of nodes: the 18 TCP-AR bipolar EEG channels, five cortical regions (frontal, temporal, central, parietal, and occipital), and three hemispheric groups (left, right, and midline). These nodes are interconnected through five clinically meaningful relationship types, including channel-to-region associations, channel-to-hemisphere assignments, spatial adjacency between channels sharing common electrodes, contralateral correspondences between anatomically symmetric channel pairs, and anatomical connections among neighboring cortical regions. To further embed clinical knowledge, each channel is assigned a region-specific seizure-onset prior based on the observed prevalence of focal seizure origins across cortical regions, with the highest prior assigned to temporal regions (0.60), followed by frontal (0.20), central (0.10), parietal (0.06), and occipital (0.04) regions. The graph additionally contains electrode-level artifact susceptibility priors; however, these are not utilized in the current evaluation because TUSZ lacks artifact annotations required for validation. The overall graph design, connectivity patterns, and prior information are derived from established neurophysiological knowledge regarding EEG electrode placement, cortical organization, seizure-onset localization, artifact behavior, and the official NEDC TUSZ annotation guidelines, providing a clinically grounded foundation for graph-based seizure detection and interpretation.

### 3.7. Knowledge-Graph-Informed Graph Attention and Onset Supervision

The graph-attention component of the detector incorporates information from the EEG knowledge graph through two complementary mechanisms. First, the attention topology is partially defined by clinically derived graph relationships, including spatial channel adjacency, contralateral symmetry, and intra-regional connectivity. This clinically informed graph is combined with a data-driven functional connectivity graph generated from window-level inter-channel correlations, enabling the model to leverage both anatomical knowledge and dynamic signal interactions during message passing. Second, channel-specific seizure-onset and artifact-related priors are appended to the temporal convolutional embeddings as additional node attributes, allowing the graph-attention network to directly utilize clinically relevant localization information when learning channel representations.

To enhance sensitivity to seizure initiation patterns, an auxiliary onset-channel prediction head is introduced alongside the primary seizure detection branch. This module operates on the shared graph-attention features and is trained using a focal-masked, class-balanced channel-level binary cross-entropy loss to identify expert-annotated seizure-onset channels. The onset-localization branch functions independently of the seizure classification output and does not directly influence the final seizure probability score. Instead, it is optimized jointly with the seizure detector within a multi-task learning framework, allowing onset-channel supervision to shape the shared feature representations and improve the spatial awareness of the graph-attention layers. Importantly, model selection for the detector is based exclusively on window-level AUROC measured on the development set. Although onset annotations are used as auxiliary supervision during training, they play no role in early stopping, threshold optimization, or any model-selection procedures related to seizure detection performance.

## 4. ALARM-Net Method

### 4.1. Motivation

A straightforward strategy for converting window-level seizure probabilities into seizure events is to apply a probability threshold and merge adjacent positive windows into continuous alarm segments. However, when applied to TUSZ, this approach results in a highly skewed distribution of false alarms across subjects. A small subset of patients—approximately the worst-performing 10% of the cohort (four subjects)—contributes a disproportionately large fraction of the overall false-alarm burden. For these subjects, the average false alarms per 24 hours (FA/24h) reaches several tens on both the development and evaluation sets. In contrast, the median subject in the evaluation set experiences no false alarms, while the median subject in the development set still exhibits a noticeable level of false detections.

These findings highlight that false alarms are concentrated within a relatively small group of challenging patients and motivate the need for an event-level mitigation strategy. Rather than redesigning or replacing the underlying seizure detector, ALARM-Net addresses this problem by operating on the temporal and structural characteristics of generated alarm candidates. By analyzing alarm proposals after window-level classification, the framework aims to identify and suppress clinically irrelevant false positives that persist despite strong window-level detection performance, thereby improving the practical utility of automated seizure monitoring systems.

### 4.2. Pipeline

ALARM-Net decomposes the seizure alarming process into two clinically complementary modules (Figure 1). The first is a knowledge-graph-informed, onset-aware seizure detector that interprets multichannel EEG activity within a clinically grounded anatomical framework and incorporates seizure-onset localization information (Sections 3.6–3.7). The second is KGSuppressor, a knowledge-graph-driven event-level suppression module that transforms the detector’s probability outputs into clinically actionable alarm events. This modular design addresses two key clinical objectives: ensuring that the detector’s spatial reasoning aligns with anatomically plausible seizure-onset patterns, and reducing the false-alarm burden of the generated alarm stream while maintaining predefined sensitivity-loss constraints.

**Figure 1.**
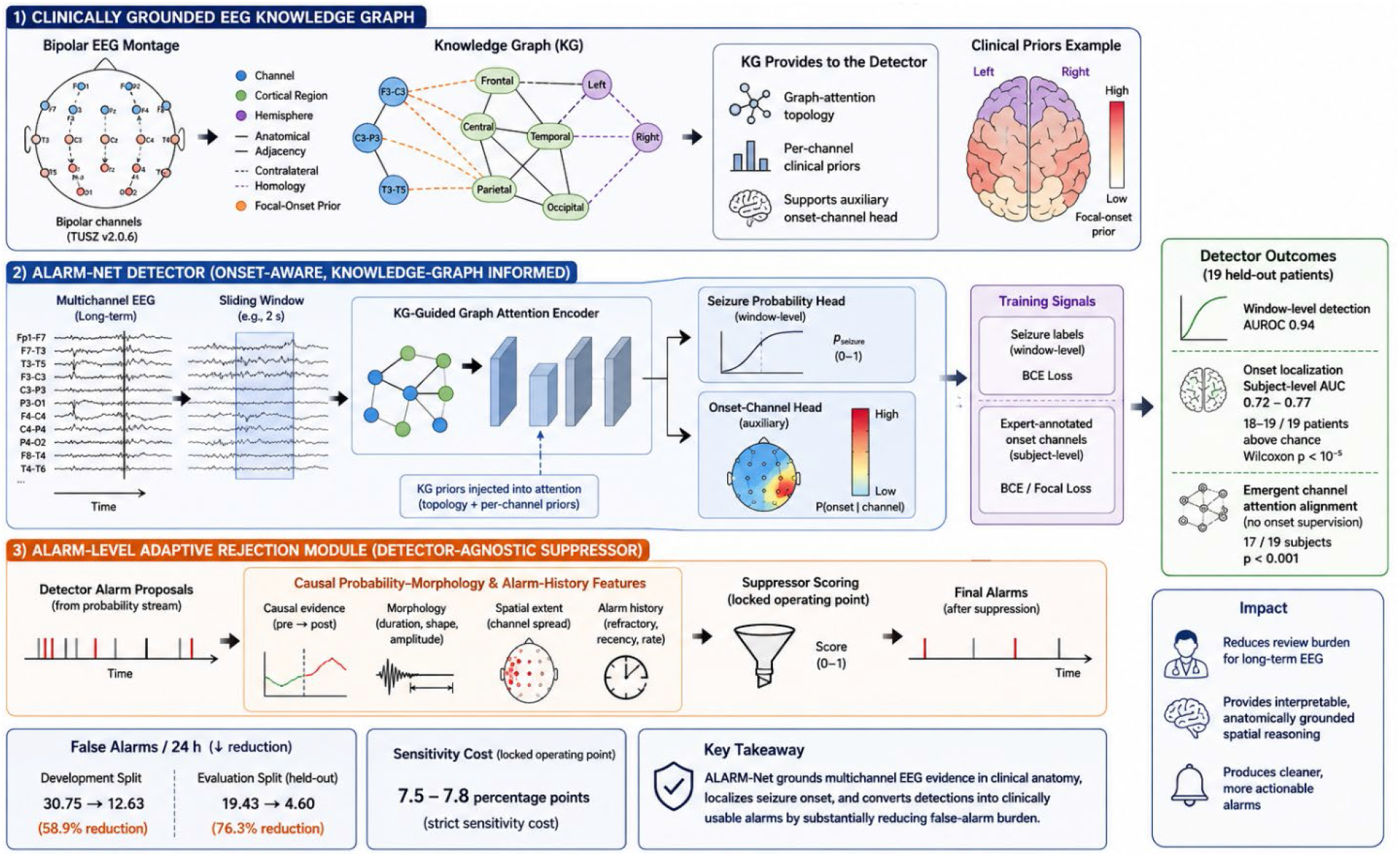
Overview of the proposed ALARM-Net framework for knowledge graph-informed, onset-aware multichannel EEG seizure detection and event-level false-alarm suppression.

In the current study, the suppression module is applied to the fixed probability outputs generated by Run A, allowing the contribution of event-level alarm suppression to be evaluated independently of the seizure detector itself. Consequently, KGSuppressor represents the only trainable component within the suppression stage. The knowledge-graph-informed detector, meanwhile, is assessed separately with respect to its seizure detection performance, preservation of sensitivity, and ability to provide clinically meaningful onset localization and anatomical interpretability (Section 5.8).

#### Stage 1: Alarm Proposal Generation

The first stage of ALARM-Net generates high-recall seizure alarm candidates from the detector’s probability output stream. For each EEG recording, window-level seizure probabilities are first transformed into a continuous per-second probability sequence by averaging predictions from all overlapping windows covering each time point. This probability timeline is then smoothed using a 60-second moving average filter to reduce short-term fluctuations. Time points whose smoothed probabilities exceed a predefined threshold (τ_p_ = 0.10) are marked as potential seizure activity, and consecutive positive intervals are grouped into candidate alarm events. To account for fragmented detections, candidate events separated by less than 120 seconds (τg) are merged into a single proposal, while events shorter than 10 seconds (τd) are discarded. These parameters are optimized on the development set to maximize event-level recall while maintaining a permissive operating regime. Consequently, this stage is intentionally designed to generate a broad set of alarm proposals, prioritizing sensitivity over precision.

#### Stage 2: Knowledge-Graph-Informed Alarm Suppression

In the second stage, each alarm proposal is evaluated using the proposed KGSuppressor framework. For a given proposal, channel-level feature embeddings generated by the detector’s TCN-GAT encoder are aggregated over the proposal duration, producing a representation for each of the 18 EEG channels. These channel embeddings are assigned to the nodes of a proposal-specific heterogeneous graph whose structure follows the clinically informed EEG knowledge graph described in Section 3.6. The KGSuppressor performs relational message passing across channels, cortical regions, and hemispheric structures to estimate the probability that a proposal corresponds to a true seizure event rather than a false alarm.

For comparison, a tabular baseline model is constructed using 14 causal probability-morphology features, all of which can be computed using information available up to the proposal endpoint. These include ten probability-shape descriptors (maximum, mean, median, standard deviation, duration, probability-area-under-the-curve, start and end probabilities, linear probability trend, and peak count), one local contextual feature representing the average probability during the preceding 60 seconds, and three alarm-history features characterizing previous alarm occurrences within the same recording. Both suppression approaches operate causally, relying only on information available before the proposal decision time. The final suppression decision is made after the merge-gap interval has elapsed. In the proposed framework, KGSuppressor assigns each proposal a retention probability, and alarms exceeding a decision threshold (τs) are preserved as final seizure alarms.

### 4.3. Training Labels

For each alarm proposal generated during training, a binary label is assigned based on its temporal overlap with annotated ground-truth seizure events. A proposal is labeled as positive (label = 1) if it overlaps with at least one ground-truth seizure event for 10 seconds or longer; otherwise, it is labeled as negative (label = 0). This overlap criterion is intentionally more stringent than conventional any-overlap labeling approaches, as it reflects a minimum duration required for an alarm to provide clinically meaningful warning and intervention value. The impact of this labeling strategy is further investigated through an ablation study in Section 5.6, where the proposed strict-overlap criterion is compared against a more permissive any-overlap labeling scheme.

### 4.4. Suppressor Backbone

KGSuppressor is a heterogeneous graph neural network designed for event-level false-alarm suppression by leveraging channel-level embeddings extracted from the TCN-GAT seizure detector and organizing them according to a clinically informed EEG knowledge graph. For each alarm proposal, channel embeddings are aggregated and mapped onto a heterogeneous graph consisting of channel, cortical-region, and hemisphere nodes connected through anatomical and functional relationships. The model employs two heterogeneous graph convolution layers with relational message passing, followed by graph pooling and a multilayer perceptron to estimate the likelihood that an alarm corresponds to a true seizure event. Training is performed using AdamW optimization, class-balanced binary cross-entropy loss, early stopping, and ensemble averaging across multiple random seeds.

The use of KGSuppressor is motivated by its ability to exploit clinically meaningful graph structure while preserving rich spatial information contained in channel-level detector embeddings. Ablation studies demonstrate the importance of the knowledge-graph topology compared with non-structured graph variants. Experimental results show that KGSuppressor achieves substantially better sensitivity preservation than conventional tabular approaches, reducing false alarms while incurring only a 0.8 percentage-point sensitivity loss, compared with 7.8 percentage points for a CatBoost-based baseline. Owing to its superior performance, clinically grounded reasoning, and seamless integration with the graph-based seizure detection framework, KGSuppressor was adopted as the primary event-level suppression method. The entire framework was implemented in PyTorch [24] and PyTorch Geometric [25] and trained on an NVIDIA A100 GPU.

### 4.5. Excluded Features

KGSuppressor operates directly on channel-level detector embeddings, eliminating the need for manual feature engineering and handcrafted alarm descriptors. The 14 causal probability-morphology features described in Section 4.2 are retained solely for comparison with a gradient-boosting baseline (Section 5.5). During feature selection for this baseline, two additional candidate features were evaluated but ultimately excluded. The first, recording_duration_s, represents recording-level context rather than alarm-specific evidence and was found to bias the model toward recording-length-dependent decisions, reducing generalizability across datasets with different duration distributions. Ablation experiments (Section 5.6) further showed that this feature dominated feature-importance rankings, overshadowing clinically relevant event-morphology information. The second, post60_mean_p, captures the average probability during the 60 seconds following an alarm proposal and was excluded because it relies on future information, making it unsuitable for causal, real-time deployment. Its removal preserved a fully causal inference framework without degrading performance.

### 4.6. Operating-Point Selection

The suppression threshold (τ_s_) for KGSuppressor was determined on the development set using clinically motivated operating constraints defined relative to the detector-only baseline. Specifically, threshold selection was required to satisfy two criteria: (1) a strict sensitivity reduction of no more than 8 percentage points, and (2) a false-alarm rate not exceeding 12 FA/24h on the development set. The FA/24h constraint directly limits the absolute alarm burden, ensuring that the resulting alarm rate remains clinically manageable regardless of recording duration.

Among all thresholds satisfying these requirements, the final operating point was chosen as the one yielding the highest strict sensitivity while maintaining FA/24h ≤ 12.0. Once selected on the development set, this threshold was fixed and applied unchanged to the held-out evaluation set. An equivalent threshold-selection procedure was independently performed for the tabular baseline model.

### 4.7. Evaluation Protocol

Event-level evaluation is performed using a one-to-one matching strategy with a 30-second temporal collar, allowing for uncertainty in seizure onset annotations. A predicted alarm is considered a true positive if its expanded interval overlaps an unmatched ground-truth seizure event, while each ground-truth event can be matched only once. The false alarms per 24 hours (FA/24h) metric is calculated as the total number of false-positive alarms divided by the cumulative non-seizure recording time and normalized to a 24-hour period, providing a duration-weighted measure of alarm burden across the cohort. Separate per-subject false-alarm statistics are also reported using an unweighted definition.

Both the tabular baseline and KGSuppressor operate under a causal inference framework, where all features and channel embeddings are computed using information available up to the end of each alarm proposal. However, final alarm confirmation requires waiting for the merge-gap interval (τg = 120 s) to determine whether additional high-probability segments should be merged with the current proposal. Consequently, the system introduces a fixed and predictable alarm-finalization latency determined by the proposal-generation process. This merge-gap delay represents the primary contributor to alarm-to-clinician latency in prospective deployment and reflects a trade-off between timely notification and robust event aggregation.

## 5. Results

### 5.1. Main Comparison — Dev

Table 1 presents the event-level performance on the development set, comparing the detector-only baseline with the proposed KGSuppressor operating at the development-set-selected threshold (τ_s_ = 0.50). The results show that KGSuppressor decreases FA/24h by 29.5% while incurring only a 2.5 percentage-point reduction in strict sensitivity. Although the improvement in F1 score is relatively small (+0.031), the primary clinical benefit lies in the suppressor’s ability to maintain seizure detection performance while substantially reducing false alarms. Compared with alternative suppression strategies operating at similar false-alarm rates, KGSuppressor preserves a greater proportion of true seizure detections. Furthermore, the median seizure-onset detection timing remains unchanged relative to ground-truth annotations, indicating that the suppression process reduces false alarms without introducing additional delays in alarm generation.

**Table 1.** Dev-split event-level metrics (Run A baseline vs KGSuppressor at the dev-selected operating point). All metrics strict (denominator = all 321 GT events).

| **Metric** | **Detector-only** | **KGSuppressor** | **$\Delta$** |
| --- | --- | --- | --- |
| Strict F1 | 0.435 | **0.466** | **+0.031** |
| Strict sensitivity | 0.502 | 0.477 | −2.5 pp |
| Precision | 0.384 | **0.457** | **+0.073** |
| FA/24h | 16.86 | **11.89** | **−29.5%** |
| Onset delay (median, s) | −45.8 | −56.0 | −10.2 s |

### 5.2. Held-Out Eval Performance

The operating configuration determined on the development set—comprising the proposal-generation parameters (τ_p_, τg, τd) = (0.10, 120, 10) and a suppression threshold of τ_s_ = 0.50—was fixed and applied directly to the held-out evaluation set without any additional tuning. The resulting event-level performance is reported in Table 2.

**Table 2.** Eval-split event-level metrics (held-out test, dev-locked operating point). All metrics strict (denominator = all 360 GT events).

| **Metric** | **Detector-only** | **KGSuppressor** | **$\Delta$** |
| --- | --- | --- | --- |
| Strict F1 | 0.589 | **0.625** | **+0.036** |
| Strict sensitivity | 0.525 | 0.517 | −0.8 pp |
| Precision | 0.670 | **0.791** | **+0.121** |
| FA/24h | 19.55 | **10.30** | **−47.3%** |
| Onset delay (median, s) | −37.0 | −39.0 | −2 s |

Two key findings emerge from this evaluation. First, the detector-only baseline exhibits a higher false-alarm burden on the evaluation set than on the development set (19.55 vs. 16.86 FA/24h), indicating differences in alarm characteristics between the two cohorts. Second, the benefits of ALARM-Net generalize well to the unseen evaluation data and remain consistent with those observed on the development set. Specifically, the reduction in strict sensitivity is limited to 2.5 percentage points on the development set and 0.8 percentage points on the evaluation set, demonstrating stable sensitivity preservation across splits. At the same time, ALARM-Net achieves substantial reductions in false alarms, decreasing FA/24h by 29.5% on the development set and 47.3% on the evaluation set, while improving precision by 0.073 and 0.121, respectively.

Although the gain in strict F1 score is relatively modest (+0.031 on development and +0.036 on evaluation), this is partly attributable to the already strong detector-only baseline performance, which leaves limited room for further improvement. More importantly, the clinically meaningful outcomes—namely, significant reductions in false alarms and notable improvements in precision—are maintained and even strengthened on the held-out evaluation cohort. These results demonstrate that the operating point selected on the development set transfers successfully to the evaluation set without requiring recalibration or threshold adjustment.

### 5.3. patient-Level False-Alarm Burden

The global FA/24h values reported in Tables 1 and 2 are calculated using a duration-weighted formulation, defined as the total number of false-positive alarms across the entire cohort divided by the cumulative non-seizure recording time and normalized to a 24-hour period. In contrast, the per-subject FA/24h values presented in this section are computed individually for each subject and then summarized across subjects without weighting by recording duration. While the per-subject metric provides insight into patient-specific alarm burden, it is inherently more sensitive to subjects with shorter recordings. Consequently, global and per-subject FA/24h values represent different perspectives on alarm burden and should not be interpreted as directly comparable quantities. Per-subject analyses are restricted to subjects for whom the detector generated at least one alarm proposal at the selected operating point. Table 3 presents the resulting distribution for the detector-only baseline.

**Table 3.** Per-subject FA/24h summary statistics for the detector-only baseline on dev and eval splits, computed over subjects for which the detector produced at least one proposal (dev n = 40; eval n = 41). Worst-decile mean is the unweighted average over the four subjects (10% of cohort) with the highest per-subject FA/24h on each split. Per-subject FA/24h is computed using each subject’s own non-seizure recording duration as the denominator.

| Statistic | Dev detector ($n = 40$) | Eval detector ($n = 41$) |
| --- | --- | --- |
| Median per-subject FA/24h | 18.42 | 0 |
| Worst-decile mean | 79.55 | 52.77 |

The per-subject false-alarm distribution reveals substantial heterogeneity and exhibits a pronounced right-skewed pattern. On the evaluation set, the median subject experiences no false alarms, whereas the average false-alarm burden among subjects in the worst-performing decile exceeds 50 FA/24h. A similar trend is observed on the development set, where even the median subject experiences a measurable false-alarm rate and the worst-decile mean approaches 80 FA/24h. This concentration of false alarms among a small subset of challenging subjects provides strong motivation for incorporating an event-level suppression mechanism. Because post-suppression per-subject FA/24h estimates can be disproportionately affected by short-duration recordings—for example, a single false alarm in a 30-minute recording corresponds to approximately 48 FA/24h—the primary suppression outcome in this study is based on the global duration-weighted FA/24h metric. Patient-level summaries are therefore reported as descriptive analyses rather than primary performance outcomes. Developing more robust patient-level evaluation measures that explicitly account for recording duration remains an area for future investigation.

It is also important to note that the worst-decile statistics are derived from only four subjects in each split (approximately 10% of the 40–41 subjects available). Accordingly, these values should be viewed as descriptive indicators of extreme alarm burden rather than precise population-level estimates. To provide a more balanced characterization of the distribution, both median and worst-decile statistics are reported, allowing readers to assess the spread of patient-level false-alarm burden without relying on a single summary measure.

### 5.4. Suppressor Backbone Ablation

Table 4 presents the results of the graph-structure ablation study together with comparisons against alternative suppression approaches on the held-out evaluation set using operating points determined exclusively on the development set. The full KGSuppressor, which incorporates the complete EEG knowledge graph, achieves a strict sensitivity of 0.517, representing only a 0.8 percentage-point reduction relative to the detector-only baseline sensitivity of 0.525. This performance is obtained at 10.30 FA/24h, with a corresponding F1 score of 0.625 and precision of 0.791. In contrast, the fully connected graph variant, where all channel nodes are interconnected and the clinically informed knowledge-graph relationships are removed, achieves a lower strict sensitivity of 0.506 (a 1.9 percentage-point reduction from baseline) while operating at the same false-alarm rate of 10.30 FA/24h, yielding an F1 score of 0.616. The additional 1.1 percentage-point sensitivity preservation achieved by the full-KG model suggests that the clinically derived graph topology contributes meaningful relational information for distinguishing true seizure events from false alarms beyond what can be captured through unrestricted channel connectivity alone.

**Table 4.** Graph-structure ablation and baseline comparison (eval, at dev-selected operating points).

| **Suppressor** | **Strict F1** | **Strict sens** | **Precision** | **FA/24h** |
| --- | --- | --- | --- | --- |
| Detector-only | 0.589 | 0.525 | 0.670 | 19.55 |
| KGSuppressor | 0.625 | 0.517 | 0.791 | 10.30 |
| KGSuppressor<br>(fully-connected) | 0.616 | 0.506 | 0.788 | 10.30 |
| CatBoost<br>(baseline) | 0.560 | 0.447 | 0.749 | 11.35 |

A comparison with the CatBoost gradient-boosting baseline further highlights the benefits of graph-based event-level reasoning. Using the 14 causal probability-morphology features, CatBoost achieves a strict sensitivity of 0.447, corresponding to a 7.8 percentage-point decline from the detector-only baseline, at 11.35 FA/24h, with an F1 score of 0.560. Relative to this tabular approach, KGSuppressor preserves approximately 7.0 percentage points more sensitivity while operating at a comparable or lower false-alarm rate. These findings demonstrate that exploiting channel-level detector embeddings and clinically informed spatial relationships through graph neural networks provides a substantial advantage over conventional tabular classification methods for event-level false-alarm suppression.

### 5.5. Rule-Based Baselines

To assess whether simple and interpretable suppression strategies could provide performance comparable to KGSuppressor, four rule-based alarm-filtering methods were evaluated using the same set of alarm proposals. These rules retained alarms based on: (R1) a threshold on the maximum proposal probability (max_p), (R2) a threshold on proposal duration (duration_s), (R3) a threshold on the mean probability during the 60 seconds preceding the proposal (pre60_mean_p), and (R4) a combination of these criteria requiring high maximum probability, sufficient duration, and low pre-alarm probability. Each rule was optimized on the development set under a sensitivity-loss constraint and subsequently evaluated on the held-out test set using the same protocol applied to KGSuppressor.

The results indicate that simple threshold-based rules are unable to match the performance of the graph-based suppressor. Rules R1, R2, and R4 were able to approach the development-set clinical operating constraints, whereas R3 failed to satisfy the false-alarm budget and effectively converged to a no-suppression solution. Under the selected constraints, R4 reduced to a single-variable threshold and therefore produced identical operating points to R1. Although R1 and R4 achieved false-alarm rates close to the target budget, they incurred a 7.5 percentage-point sensitivity reduction, while R2 preserved more sensitivity but at the expense of substantially higher false-alarm rates. When transferred to the held-out evaluation cohort, KGSuppressor demonstrated markedly better sensitivity preservation, exhibiting only a 0.8 percentage-point decrease in strict sensitivity, compared with 7.8 percentage points for the CatBoost baseline and 12.2 percentage points for the best rule-based method (R1). These findings suggest that exploiting channel-level spatial representations and clinically informed graph relationships provides a substantial advantage over both simple threshold rules and conventional tabular classifiers for event-level false-alarm suppression. Nevertheless, broader validation on additional EEG datasets would be necessary to establish the generalizability of these observations.

### 5.6. Configuration A → F Ablation

To further investigate the impact of design choices in the CatBoost tabular baseline, we evaluated multiple configurations that varied the inclusion of recording-duration information, post-proposal features, labeling strategies, and regularization settings. The original ablation study focused primarily on assessing the effects of removing recording_duration_s and modifying the proposallabeling criterion. It is important to note that these design considerations apply exclusively to the CatBoost baseline and serve as supplementary diagnostic analyses; they do not affect KGSuppressor, which operates directly on channel-level detector embeddings and does not depend on handcrafted feature engineering or manual feature-selection decisions. The evaluated configurations and their corresponding development-set operating points are summarized in Table 6.

The results of the rerun confirm that Configuration F corresponds to the CatBoost model used in the primary comparison reported in Table 5 and is therefore retained as the reference tabular baseline. Overall, the A–F configurations highlight the substantial sensitivity of tabular classifiers to feature-selection choices, labeling definitions, and regularization strategies. Among the evaluated alternatives, Configuration F—combining strict proposal labeling, exclusion of selected features, and regularization—provides the strongest tabular performance. Nevertheless, none of the CatBoost configurations achieve sensitivity preservation comparable to that of KGSuppressor, further emphasizing the advantages of graph-based event-level reasoning over conventional feature-engineered tabular approaches.

**Table 5.** Rule-based suppression baselines compared to KGSuppressor.

| Suppressor | Split | Strict F1 | Strict sens | Precision | FA/24h |
| --- | --- | --- | --- | --- | --- |
| Detector-only | dev | 0.435 | 0.502 | 0.384 | 16.86 |
| R1 (max_p ≥ 0.30) | dev | 0.489 | 0.427 | 0.573 | 12.16 |
| R2 (duration ≥ 90s) | dev | 0.464 | 0.461 | 0.467 | 20.14 |
| R3 (pre60 ≤ 0.10) | dev | 0.435 | 0.502 | 0.384 | 16.86 |
| R4 (combined) | dev | 0.489 | 0.427 | 0.573 | 12.16 |
| CatBoost (baseline) | dev | 0.486 | 0.427 | 0.564 | 11.44 |
| Detector-only | eval | 0.589 | 0.525 | 0.670 | 19.55 |
| R1 (max_p ≥ 0.30) | eval | 0.552 | 0.403 | 0.879 | 4.18 |
| R2 (duration ≥ 90s) | eval | 0.584 | 0.481 | 0.746 | 12.33 |
| R3 (pre60 ≤ 0.10) | eval | 0.590 | 0.503 | 0.713 | 15.25 |
| R4 (combined) | eval | 0.552 | 0.403 | 0.879 | 4.18 |
| CatBoost (baseline) | eval | 0.560 | 0.447 | 0.749 | 11.35 |

**Table 6.** Supplementary tabular-baseline ablation on dev (CatBoost only; not used for KGSuppressor selection). Each row reports the best-strict-F1 dev operating point for that configuration. These configurations characterize the tabular CatBoost baseline only and are not used for KGSuppressor selection or headline claims. Strict F1 here uses the same denominator (321 dev GT events) as Tables 1 and 2 to ensure consistency.

| Config | Duration feature | post60 | Label | Regularized | Strict F1 | FA/24h | Strict sens drop |
| --- | --- | --- | --- | --- | --- | --- | --- |
| A. baseline | yes | yes | any-overlap | no | 0.450 | 16.80 | −8.4 pp |
| B. drop duration | no | yes | any-overlap | no | 0.486 | 18.59 | −2.5 pp |
| C. strict label (overlap $\geq 10$s) | yes | yes | strict_s | no | 0.452 | 22.88 | −3.4 pp |
| D. strict label (overlap_frac $\geq 0.20$) | yes | yes | strict_f | no | 0.435 | 16.86 | 0.0 pp |
| E. regularized baseline | yes | yes | any-overlap | yes | 0.457 | 10.37 | −12.5 pp |
| **F. CLEAN (tabular baseline)** | **no** | **no** | **strict_s** | **yes** | **0.486** | **12.63** | **−7.5 pp** |

### 5.7. Knowledge-Graph-Grounded Attention Validation

To evaluate the clinical relevance of the detector’s learned spatial representations, we mapped the Graph Attention Network (GAT) channel-attention weights onto the anatomical structure encoded by the EEG knowledge graph and compared them with expert-annotated seizure-onset channels extracted from the term-based TUSZ annotations. These onset annotations were not incorporated into model training, threshold optimization, or model-selection procedures and were used solely for independent post-hoc validation. Analysis of 173 focal seizure events from 19 held-out evaluation subjects demonstrated that the detector’s channel-attention patterns aligned with clinically annotated seizure-onset regions at levels exceeding chance performance, achieving a mean event-level channel AUC of 0.67, with 86% of seizure events exhibiting above-chance alignment.

This relationship remained consistent when evaluated at the subject level, which provides a more appropriate statistical unit because of within-subject event correlations. Specifically, 17 of 19 subjects demonstrated above-chance average alignment, yielding a mean subject-level AUC of 0.69 and a statistically significant Wilcoxon signed-rank test result (p < 0.001). The strongest correspondence was observed for focal seizures with highly localized onset patterns involving three or fewer onset channels (AUC = 0.77), whereas broader onset patterns showed weaker alignment (AUC = 0.62). Notably, the two subjects exhibiting below-chance alignment were both characterized by diffuse seizure onset involving eight annotated onset channels. Although the observed alignment is moderate rather than definitive and attention weights should not be interpreted as direct localization outputs, the consistent above-chance correspondence suggests that the detector’s spatial attention is meaningfully associated with clinically relevant seizure-onset regions. These findings support the role of the EEG knowledge graph in guiding anatomically grounded and interpretable spatial reasoning within the detection framework (Table 7).

**Table 7.** Knowledge-graph-grounded attention-onset alignment on held-out focal seizures. Channel-level AUC quantifies agreement between detector attention and expert onset channels; p-values are from Wilcoxon signed-rank tests against chance (AUC = 0.5). Onset annotations were used only for this post-hoc validation.

| Stratum | n | Mean AUC | p |
| --- | --- | --- | --- |
| All focal events | 173 | 0.67 | $< 1e-21$ |
| Subject-level | 19 | 0.69 | $3.6e-4$ |
| Narrow onset ($\leq 3$ ch) | 59 | 0.77 | $1.4e-9$ |
| Wide onset ($> 3$ ch) | 114 | 0.62 | $3.1e-14$ |

### 5.8. Knowledge-Graph-Informed Onset-Aware Detection

While Section 5.7 focused on analyzing the detector’s emergent attention patterns, this section directly evaluates the performance of the onset-supervised detector. Table 8 presents a comparison of three models trained under identical conditions: the baseline detector (Run A) without onset supervision, an onset-aware detector using an anatomical k-nearest-neighbor graph (AnatGAT), and a knowledge-graph-guided detector (KG-GAT) that incorporates the clinically informed graph structure and seizure-onset priors described in Section 3.7. All three models maintain strong seizure detection capability, achieving comparable development-set AUROC values ranging from 0.937 to 0.940.

**Table 8.** Detector variants under identical training. Detection AUROC is dev window-level; onset AUC and top-1 are subject-level means on the held-out eval split (19 patients), with both supervised variants exceeding chance in 18–19 of 19 patients (Wilcoxon p < 10−5; per-window chance ≈ 0.24). AnatGAT and KG-GAT differ only in the channel graph (anatomical k-NN vs knowledge-graph adjacency).

| Variant | Channel graph | Onset head | Detection AUROC | Onset AUC (subj) | Top-1 (subj) |
| --- | --- | --- | --- | --- | --- |
| Base (Run A) | k-NN + correlation | — | 0.937 | — | — |
| AnatGAT | k-NN + correlation | Yes | 0.938 | 0.77 | 0.57 |
| **KG-GAT** | KG + correlation | Yes | **0.940** | 0.72 | **0.59** |

On the held-out evaluation cohort, the auxiliary onset-localization branch successfully identifies expert-annotated seizure-onset channels at levels significantly above chance. Performance is reported at the subject level (19 patients) rather than the window level because seizure windows from the same subject are not statistically independent, and pooled analyses could lead to overly optimistic estimates. Both onset-supervised models demonstrated significant localization capability, exceeding chance performance in 18–19 of the 19 patients (Wilcoxon signed-rank p < 10^−5^). The AnatGAT model achieved a mean subject-level AUC of 0.77, while the KG-GAT model achieved a mean AUC of 0.72. The performance differences between the two models were small, with overlapping bootstrap confidence intervals indicating comparable predictive ability.

Importantly, although the knowledge-graph-based model achieves localization accuracy similar to the anatomical graph approach, it offers a more interpretable representation by encoding channel relationships through explicit clinical concepts such as anatomical adjacency, hemispheric homology, and seizure-onset priors rather than purely geometric proximity. The results demonstrate that both onset-supervised architectures can recover clinically meaningful seizure-onset regions in previously unseen patients. Together with the attention-alignment analysis presented in Section 5.7, these findings provide two complementary forms of evidence—implicit spatial reasoning reflected in emergent attention and explicit localization learned through onset supervision—supporting the conclusion that the detector’s spatial representations are grounded in clinically relevant seizure-onset anatomy.

## 6. Discussion

The principal contribution of ALARM-Net is its ability to significantly reduce the event-level false-alarm burden while preserving seizure detection performance. On the held-out evaluation cohort, KGSuppressor reduces FA/24h by 47.3% (19.55 → 10.30) with only a 0.8 percentage-point loss in strict sensitivity, whereas a CatBoost-based baseline achieves comparable false-alarm reduction at the cost of a substantially larger 7.8 percentage-point sensitivity decrease. Analysis of the detector-only baseline further reveals that false alarms are disproportionately concentrated among a small subset of patients, highlighting the importance of event-level suppression strategies. From a clinical standpoint, the reduction in FA/24h increases the average interval between false alarms from approximately 74 minutes to 2.3 hours, thereby reducing alarm fatigue and improving clinical usability [19–22]. The ability of KGSuppressor to achieve this improvement while maintaining nearly all seizure detections makes it particularly suitable for safety-critical clinical environments. Furthermore, the suppression threshold can be adjusted to accommodate institution-specific preferences regarding the trade-off between false-alarm reduction and sensitivity preservation.

All headline results are reported using strict event-level evaluation, where every annotated seizure event is included and unreachable events are counted as misses. Although reachability-adjusted sensitivities are higher, they are presented only as supplementary analyses because such metrics depend on method-specific definitions and may overestimate performance. Importantly, the operating point selected on the development set generalized directly to the held-out evaluation cohort without any retuning, producing consistent reductions in false alarms with minimal sensitivity loss. This stable transfer across subject-independent cohorts suggests that the knowledge-graph-guided structural prior provides robust and generalizable event-level reasoning for clinically deployable seizure alarm systems.

Comparison with tabular and rule-based suppression methods indicates that simple threshold-based approaches can achieve competitive performance on the development set but exhibit reduced robustness on unseen data. Although a single-threshold rule slightly outperformed CatBoost in terms of F1 score on the development cohort, the CatBoost baseline preserved seizure sensitivity more effectively on the evaluation set, suggesting that learned models can capture additional feature interactions that become important under distribution shifts. The results also show that false-alarm behavior is largely driven by a small number of dominant features, which simple thresholding rules can exploit, but they often fail to model more subtle patterns required for consistent generalization across cohorts.

From a deployment perspective, rule-based suppression provides a transparent and easily interpretable solution when cohort-specific tuning is feasible. However, for real-world settings where patient populations may differ from the training data and frequent recalibration is impractical, the KGSuppressor offers a more robust alternative, delivering superior sensitivity preservation and more stable performance across independent datasets. Furthermore, experiments with alternative detector architectures—including deeper TCNs, GRU-enhanced temporal models, and multi-scale TCN variants—yielded only marginal improvements in window-level performance and did not alleviate the concentration of false alarms among a small subset of patients. These findings motivated the final design choice of maintaining a fixed detector and focusing on event-level false-alarm suppression as the most effective strategy for improving clinical alarm performance.

A central innovation of ALARM-Net is the EEG knowledge graph, which encodes clinically meaningful relationships such as anatomical adjacency, hemispheric homology, and seizure-onset priors to provide a transparent and interpretable foundation for spatial reasoning. Rather than serving solely as a performance-enhancement mechanism, the knowledge graph enables the detector’s decisions to be expressed in clinically understandable terms, supporting seizure-onset localization, anatomically grounded interpretation, and auditability. The knowledge-graph-informed detector preserves strong seizure detection performance while improving interpretability compared with purely data-driven or geometric graph approaches, and it establishes a foundation for future evidence-linked and dynamically evolving neurophysiological knowledge-graph systems.

The study also explored a neuro-symbolic conditional random field (NS-CRF) suppressor that combined structured state transitions with interpretable rules. Although this approach reduced false alarms, it resulted in excessive sensitivity loss and failed to satisfy the predefined operating constraints, leading to its exclusion from the final framework. Nevertheless, the findings suggest that future hybrid neuro-symbolic suppression models, integrating interpretable clinical knowledge with differentiable learning mechanisms, may offer a promising direction for further improving event-level seizure alarm management.

## 7. Limitations and Future Work

All experiments were conducted using the TUSZ v2.0.6 dataset, and additional validation on independent EEG seizure corpora such as CHB-MIT, Siena, and Dianalund [4,16–18] would be necessary to establish broader generalizability. The development and evaluation cohorts include only 40 and 41 subjects, respectively, for whom the detector generated alarm proposals, meaning that patient-level statistics—particularly worst-decile estimates based on four subjects—should be interpreted as descriptive indicators of alarm burden rather than population-level estimates. Although KGSuppressor achieves substantial false-alarm reduction with only a 0.8 percentage-point sensitivity loss, more aggressive suppression settings are possible at the expense of reduced sensitivity, while the CatBoost baseline offers an alternative operating point with greater sensitivity degradation.

Several methodological limitations also remain. The detector’s fixed 5-minute chunk architecture prevents inference on recordings shorter than five minutes, and the 120-second merge-gap interval introduces a deterministic alarm-finalization delay that may limit applicability in low-latency streaming environments. In addition, TUSZ annotations contain known intra- and inter-rater variability [1], and although the 30-second event-matching collar partially mitigates onset uncertainty, annotation inconsistencies may still influence performance estimates. Future work should therefore explore shorter-context detectors, streaming-friendly proposal-generation mechanisms, and reduced-latency alarm finalization strategies.

Promising future research directions include cross-corpus validation, integration with advanced EEG foundation-model encoders, and prospective deployment studies. Evaluating whether stronger window-level detectors further reduce patient-specific false alarms—or whether ALARM-Net remains largely detector-independent—would provide valuable insight into the framework’s generality. Additional opportunities include the development of soft neuro-symbolic suppression models, incorporation of dynamically updated clinical knowledge graphs, and prospective hospital-based validation studies. Together, these directions would help transition ALARM-Net from a retrospective evaluation framework to a clinically deployable seizure alarm system.

## 8. Conclusion

This study presented ALARM-Net, a knowledge-graph-informed framework for EEG seizure detection and event-level false-alarm suppression on the TUSZ v2.0.6 dataset. The framework integrates a knowledge-graph-guided graph attention detector with KGSuppressor, a heterogeneous graph neural network that leverages the same EEG knowledge graph to refine seizure alarms. Experimental results demonstrate that KGSuppressor reduces the false-alarm burden by 47.3% (19.55 → 10.30 FA/24h) while incurring only a 0.8 percentage-point loss in strict sensitivity. Ablation studies further validate the importance of the EEG knowledge graph, showing that the full graph-based suppressor preserves sensitivity better than both a fully connected graph variant (+1.1 percentage points) and a CatBoost baseline (+7.0 percentage points) at comparable false-alarm rates.

Beyond false-alarm suppression, ALARM-Net introduces an onset-aware detection architecture capable of localizing seizure-onset channels on unseen patients (subject-level AUC 0.72–0.77, with 18–19 of 19 patients performing above chance) while demonstrating alignment between learned attention patterns and clinically annotated onset regions. By encoding anatomical relationships, hemispheric homology, and seizure-onset priors, the EEG knowledge graph provides a clinically interpretable framework that makes the detector’s spatial reasoning transparent and auditable. Overall, the findings highlight the value of knowledge-graph-guided, onset-aware, and event-level reasoning for improving the clinical usability, interpretability, and deployability of automated EEG seizure monitoring systems.

## Data Availability

The Temple University Hospital EEG Seizure Corpus v2.0.6 (TUSZ) used in this study is publicly available from the Neural Engineering Data Consortium at https://isip.piconepress.com/projects/tuh_eeg/ under the TUH EEG Corpus Data Use Agreement. Code, configuration files, and paper-relevant outputs (proposal CSVs, trained suppressor model, evaluation tables) supporting this study are available at https://github.com/osmyildiz/alarm-net-tusz.

https://github.com/osmyildiz/alarm-net-tusz

## Author Contributions

Conceptualization, O.Y. and A.S.; methodology, O.Y.; software, O.Y.; validation, O.Y.; formal analysis, O.Y.; investigation, O.Y.; data curation, O.Y.; writing—original draft preparation, O.Y.; writing—review and editing, O.Y. and A.S.; visualization, O.Y.; supervision, A.S.; project administration, A.S. All authors have read and agreed to the published version of the manuscript.

## Funding

This research received no external funding.

## Institutional Review Board Statement

Not applicable. This study used the publicly available, de-identified Temple University Hospital Seizure Corpus (TUSZ) v2.0.6.

## Informed Consent Statement

Not applicable.

## Data Availability Statement

The TUSZ v2.0.6 corpus is publicly available from the Neural Engineering Data Consortium at https://isip.piconepress.com/projects/tuh_eeg/. The code, configuration files, and paper-relevant outputs (proposal CSVs, trained suppressor model, evaluation tables) supporting this study will be made publicly available at https://github.com/osmyildiz/alarm-net-tusz upon acceptance, and are available to reviewers upon request during peer review.

## Acknowledgments

Computations were performed on the DGX A100 cluster at the University at Albany, SUNY. During the preparation of this manuscript, the authors used generative AI tools for language editing. The authors reviewed and edited the output and take full responsibility for the content of the publication.

## Conflicts of Interest

The authors declare no conflicts of interest.

## Notes

### Competing Interest Statement

The authors have declared no competing interest.

### Funding Statement

This study did not receive any external funding. Computations were performed on the DGX A100 cluster at the University at Albany, State University of New York.

### Author Declarations

Ethics committee/IRB of Temple University, Philadelphia, PA, USA gave ethical approval for the original collection and public release of the Temple University Hospital EEG Seizure Corpus (TUSZ v2.0.6). The present study is a secondary computational analysis of the de-identified, publicly released TUSZ corpus, conducted under the terms of the TUH EEG Corpus Data Use Agreement. No new human subjects were recruited and no identifiable patient information was accessed.

### Summary of Updates

Summary of revision (v2) This revision substantially extends ALARM-Net by adding a knowledge-graph- grounded, onset-aware detector and a graph-based event-level suppressor. The original contribution (event-level false-alarm suppression on TUSZ v2.0.6) is preserved; the detector and suppressor are redesigned, and additional clinical evaluation axes are introduced. 1. Title and framing. Updated from "An Event-Level False-Alarm Suppression Framework for Clinical EEG Seizure Detection" to "Knowledge Graph-Informed, Onset-Aware Multichannel EEG Seizure Detection with Event-Level False-Alarm Suppression" to reflect the new KG-grounded and onset-aware components. 2. New: clinically grounded EEG knowledge graph. v2 introduces a knowledge graph over bipolar-montage channels, cortical regions, hemispheres, anatomical adjacency, contralateral homology, and focal-onset priors. The KG is used as a computational substrate (defining graph-attention topology and per-channel clinical priors), not as a post-hoc label map. v1 did not contain a knowledge graph. 3. Detector redesign. v1 used a TCN+GAT window-level baseline ("Run A") evaluated only at the window level. v2 uses a KG-informed graph-attention detector with an auxiliary onset-channel head trained against expert- annotated seizure-onset channels; window-level AUROC is preserved (0.94). 4. New: subject-level seizure-onset localization on 19 held-out patients (subject-level AUC 0.72-0.77; 18-19 of 19 above chance; Wilcoxon p < 0.00001). v1 did not evaluate onset localization. 5. New: emergent channel-attention alignment with expert annotations in 17 of 19 subjects (p < 0.001), obtained without onset supervision. This analysis is new in v2. 6. Suppressor redesign. v1 used a CatBoost suppressor over 14 hand-crafted features. v2 introduces KGSuppressor, a knowledge-graph-informed heterogeneous GNN that scores alarm proposals using per-channel detector embeddings organized by the EEG knowledge graph. Held-out eval: FA per 24 h reduced from 19.55 to 10.30 (a 47.3% reduction) at a strict-sensitivity cost of 0.8 percentage points. The v1 CatBoost suppressor is retained as a gradient-boosting baseline, which requires 7.8 percentage points of sensitivity loss for a comparable FA reduction. 7. Editorial and compliance. Abstract reformatted for ASCII compliance (em-dash, en-dash, minus sign, and arrow characters replaced with ASCII equivalents). TUSZ version stated explicitly as v2.0.6 throughout. Unchanged: dataset (TUSZ v2.0.6), authorship, affiliations, funding declaration, competing interests, ethics statement.

